# Spatio-temporal detection for dengue outbreaks in the Central Region of Malaysia using climatic drivers at mesoscale and synoptic scale

**DOI:** 10.1101/2021.09.22.21263997

**Authors:** Stan Yip, Norziha Che Him, Nur Izzah Jamil, Daihai He, Sujit K. Sahu

**Affiliations:** Department of Applied Mathematics, The Hong Kong Polytechnic University, Hong Kong; Department of Mathematics and Statistics, Faculty of Applied Sciences and Technology, Universiti Tun Hussein Onn Malaysia, Pagoh, Johor, Malaysia; Faculty of Computer and Mathematical Sciences, Universiti Teknologi MARA, Negeri Sembilan branch, Rembau campus, Malaysia; School of Mathematical Sciences, University of Southampton, Southampton, UK

**Author notes:** Corresponding Author. Address: Department of Applied Mathematics, The Hong Kong Polytechnic University, Hung Hom, Hong Kong.

**Keywords:** Bayesian spatio-temporal model, BYM2, dengue, early warning system, ENSO, El Niño Modoki

## Abstract

The disease dengue is associated with both mesoscale and synoptic scale meteorology. Previous studies for south-east Asia have found very limited association between synoptic variables and the reported dengue cases. It will immensely beneficial to establish more clear association with rate of cases and the most relevant meteorological variables in order to institute an early warning system.

A rigorous Bayesian modelling framework is provided to identify the most important co-variates and their lagged effects for developing an early warning system in the Central Region of Malaysia.

Along with other mesoscale environmental measurements, we also examine multiple synoptic scale Niño indices which are related to the phenomenon of El Niño Southern Oscillation and an unobserved variable derived from reanalysis data. A probabilistic early warning system is built based on a Bayesian spatio-temporal hierarchical model.

Our study finds a 46.87% of increase in dengue cases due to one degree increase in the central equatorial Pacific sea surface temperature with a lag time of six weeks. We discover the existence of a mild association between the rate of cases and a distant lagged cooling effect related to a phenomenon called El Niño Modoki. These associations are assessed by using a Bayesian spatio-temporal model in terms of estimated out-of-sample predictive accuracy.

With the novel early warning system presented, our results show that the synoptic meteorological drivers can enhance short-term detection of dengue outbreaks and these can also potentially be used to provide longer-term forecasts.

**Practical Implications:** In 2019, it was reported that there is a severe dengue upsurge in Malaysia. Reported cases rose over 60% from 80,615 in the 2018 to 130,101 with 182 deaths (Rahim et al., 2021). The disease has been described as a silent killer that the infection rate once surpassed that of COVID-19. There is a need of an early warning system to alert the authority in order to identify relevant risk factors and the forthcoming outbreak hot-spots. The Bayesian hierarchical spatial dynamic model componentises different aspects of dengue dynamics into one unified model. Its flexibility allows us to regularly review the disease dynamic under changing environment and transmission mechanism such as the implementation of the Movement Control Orders (MCO) during COVID-19. Practically, this prototype model should be run at least once a week to generate forecasts which is fed with the dengue cases data from weekly press release and meteorological information from publicly available sources. By assessing the probability estimates, the alert has its intrinsic meaning and the sensitivity can be adjusted effortlessly.

**Highlights:**

- El Niño Southern Oscillation is a crucial driver to dengue outbreaks in Malaysia.
- A few different climate oscillations affect the dengue transmission pattern.
- Bayesian spatial dynamic model helps the development of early warning system.
- The model components can be added or modified under the hierarchical Bayes framework.

## 1 Introduction

Dengue is a very harmful mosquito-borne viral infection worldwide. Gubler (1998) describes that the dengue virus (DENV) is transmitted by the bite of female `Aedes aegypti` mosquitoes. To a lesser extent, `Aedes albopictus` is also a vector of indoor transmission (Noor et al., 2018). Four serotypes of virus DENV-1, DENV-2, DENV-3 and DENV-4 following the human cycle are genetically similar (Mustafa et al., 2015). This viral infectious disease which can lead to a wide spectrum of clinical manifestations such as acute onset high fever, muscle and joint pain, myalgia, cutaneous rash, hemorrhagic episodes and circulatory shock (Hasan et al., 2016). Its burden to the pubic health system is enormous. Bhatt et al. (2013) estimate there are 390 million total annual infections throughout the world. In Asia, dengue fever has been reported earliest by Skae (1902), followed by dengue hemorrhagic fever and dengue shock syndrome epidemics in the twentieth century (Henchal and Putnak, 1990).

Climate is a crucial determinant of dengue disease transmission by affecting its vector dynamics (Morin et al., 2013). Local and global climate not only influence the spatial distribution of infections (Johansson et al., 2009) but also the interanuual variability (Cazelles et al., 2005). The climatic impact to a dengue outbreak is also known to be cumulative and delayed (Lowe et al., 2018). Simple models cannot account for these complex spatial dynamic dependencies.

The Selangor state together with the two adjacent federal territories namely Kuala Lumpur and Putrajaya are collectively called the Central Region. It contributes most to the national dengue hospitalisation in Malaysia. In the region, the dengue infection rates have increased significantly in the past decade as reported by Abd Majid et al. (2021) and Salim et al. (2021). Hii et al. (2016) emphasise that dengue is a climate-sensitive infectious disease. The rapid change in climate drivers increases the risk of dengue outbreaks in the past decade. A climate-based early warning system (EWS) has the potential to enhance surveillance and control of the disease.

A significant relationship between dengue hospitalisations and covariates such as precipitation, temperature, number of monthly rain days and El Niño-Southern Oscillation (ENSO) phenomenon is found for 12 states of West Malaysia (Che Him et al., 2018b). A similar study by Che Him et al. (2018a) identifies two distinct spatial clusters via two generalised additive models (GAM) for nine districts of the state of Selangor. Ahmad et al. (2018) conduct a large scale study for 81 weeks including actively collected ovitrap and rain gauge data. A variant of linear regression model is used to identify the entomological, epidemiological and environmental drivers that contributed to the dengue outbreak of two locations in Selangor state. Salim et al. (2021) develop a supporting vector machine model that incorporates environmental variables including temperature, wind speed, humidity, and rainfall to predict dengue outbreaks.

Bayesian spatio-temporal hierarchical modelling framework for areal data (Lowe et al., 2011, 2013, 2014; Stewart-Ibarra and Lowe, 2013) is widely used in dengue disease modelling and prediction. Using spatio-temporal model as a toolkit, it can have a better capacity to handle explicit contribution from covariates information and latent spatio-temporal dependency. One popular choice of structured prior to capture spatially spill-over effect is a conditional intrinsic Gaussian autoregressive prior (CAR; Besag et al., 1991). The spatio-temporal autocorrelation, as a source of information that closer areal units and temporally close time periods tend to have more similar values (Lee et al., 2018). With the fact that spatial and temporal components are intrinsically interacted, a variety of CAR-based spatio-temporal model is developed to tackle many real-world applications as investigated by Bernardinelli et al. (1995), Knorr-Held (2000), Napier et al. (2016), Rushworth et al. (2014, 2017) and Sahu (2021).

The remainder of this paper is organised as follows. Section 2 describes the data used in this study. Section 3 illustrates the components considered in the Bayesian spatio-temporal models. The model implementation, validation, evaluation of the EWS are discussed in Section 4. A detail discussion is delivered in Section 5.

## 2 Data

### 2.1 Environmental drivers related to dengue transmission

ENSO is a global scale of climate variation that the cycles have lasted between two and seven years. Several previous studies have found there is an association between epidemic dengue and ENSO in some world populations (Kovats et al., 2003). Different regions of sea surface temperature (SST) are used to define ENSO (Rasmusson and Carpenter, 1982). Ashok et al. (2007) define the anomalous warming events that occur in the central equatorial Pacific (Niño4 region) as an alternative type of El Niño called El Niño Modoki which is different from the conventional study region Niño3.4 (5°N-5°S, 170°W-120°W). McGregor and Ebi (2018) highlight that the contrasting rainfall fields for conventional El Niño and El Niño Modoki events hint at potential spatio-temporal inconsistencies of ENSO-health associations. Salimun et al. (2014) find that, although displayed much warmer SST anomalies in the Indian Ocean and regional seas in the Maritime Continent, the impact on the winter rainfall during conventional El Niño in boreal winter season over Peninsular Malaysia is minimal but significant higher during El Niño Modoki. Tangang et al. (2017) show that, during winter, a strong La Niña leads to a significant decrease in wet precipitation extremes over the Peninsular Malaysia due to the anomalous cyclonic circulation over strong La Niña. Nevertheless, Hanley et al. (2003) demonstrate that Niño4 index is more relevant to La Niña but poorly explain El Niño whilst the Niño1+2 index has the opposite characteristics. These two SST indices altogether cover different types of ENSO and their impact on dengue transmission.

Most high impact weather in synoptic scale occurs where the atmosphere is experiencing rising motion. The vertical velocity measured by the omega equation is associated with high impact weather and cyclones (Dostalek et al., 2017). In a study of the impact of meteorological factors to the air pollution in China, Hou et al. (2018) indicate that the vertical velocity has a short-term influence on PM_2.5_ level in the Pearl River Delta. It is expected that the unobserved meteorological variable would add value to our understanding on environmental association with the disease.

Wong et al. (2011) use a lagged 22 day mean air temperature to capture the second generation gonotrophic cycle of `Aedes` mosquitoes to predict ovitrap index. Cheong et al. (2013) study the effects of temperature, rainfall and wind speed in Selangor with emphasis on their lag times. The lag times of 51 days minimum daily temperature and 28 days bi-weekly cumulated rainfall present positively associated with dengue hospitalisations. The effect from mesoscale local temperature and rainfall is related to some major other synoptic climate oscillations which influences the regional climate of Malaysia such as Indian Ocean Dipole (IOD; Tangang et al., 2012). IOD can happen in conjunction with ENSO or independently. Hong and Jin (2014) discover that the IOD-ENSO interaction is the cause of the generation of Super El Niños. Hameed et al. (2018) also show that the IOD lagged Niño3.4 by three to six months depending on location.

This study conjectures that air pollution has a profound effect on the mosquito vectors especially ozone. Thiruchelvam et al. (2018) evaluate that the relationship between air quality and dengue hospitalisations. It is asserted that the air pollution index (API) levels do not have a significant effect on the reported cases. However, ozone is proven to have a repellent effect on both `Aedes aegypti` and `Aedes albopictus` (Wan-Norafikah et al., 2016). The API used by the Malaysian government follows the Pollutant Standard Index (PSI; Swamee and Tyagi, 1999) by the United States Environmental Protection Agency (USEPA). The API is an index that the highest value of the sub-indices of five pollutants namely carbon dioxide, ozone, nitrogen dioxide, sulphur dioxide and particulate matter with a diameter of less than 10 microns taken. Its impact on humans has been thoroughly studied but its applicability to dengue transmission is questionable. Without knowing which pollutant it refers to, the lagged value of API is meaningless. For this reason, it is worthwhile to investigate individual pollutants separately.

### 2.2 Data source

The weekly counts of hospital admissions for dengue fever incidence (*Y*_*kt*_) in Selangor State and two federal territories in Malaysia (indexed *k*) from 2013 to 2019 (indexed *t*) were obtained from the Ministry of Health (MOH) Malaysia. Relevant demographic information is obtained from the Department of Statistics Malaysia (DOSM). Nine Selangor districts and two federal territories namely Kuala Lumpur and Putrajaya have been considered.

Environmental variables such as air pollution index, ozone concentration level (in part per million) and temperature are provided by the Malaysian Department of Environment (DOE), Ministry of Environment and Water whilst precipitation information (in mm) is provided by the Malaysian Meteorological Department (MetMalaysia). All ozone concentration level information in the federal territories Kuala Lumpur and Putrajaya are missing. These are imputed by the average values of their adjacent districts.

The Niño4 and Niño1+2 SST indices (Huang et al., 2021) capturing sea surface temperature anomalies in the central equatorial Pacific region (5°N-5°S, 160°E-150°W) and the region of coastal South America (0°-10°S, 90°W-80°W) are obtained from NOAA Climate Prediction Center.

Gridded (2.5° × 2.5°) reanalysis daily mean vertical velocity in pressure coordinates obtained from the NCAR/NCEP Reanalysis (Kalnay et al., 1996) is aggregated into weekly scale according to the epidemiological week (Epi week) defined by MOH.

Finally, the administrative district areal boundaries are extracted from The Humanitarian Data Exchange and all studied districts are within one (2.5° × 2.5°) grid cell in the reanalysis dataset.

### 2.3 Exploratory Data Analysis

#### 2.3.1 Basic characteristics

A total of 414284 dengue fever cases was reported in nine districts of Selangor and two federal territories from January 2013 to December 2019 in the Central Region. The total numbers of cases vary from 26422 in 2013 to 87967 in 2019. Since the outbreaks after summer in 2013, there is no clear annual trend until a severe upsurge in 2019 which surpassed three fold of the total cases in 2013 (Fig. 1).

**Figure 1:**
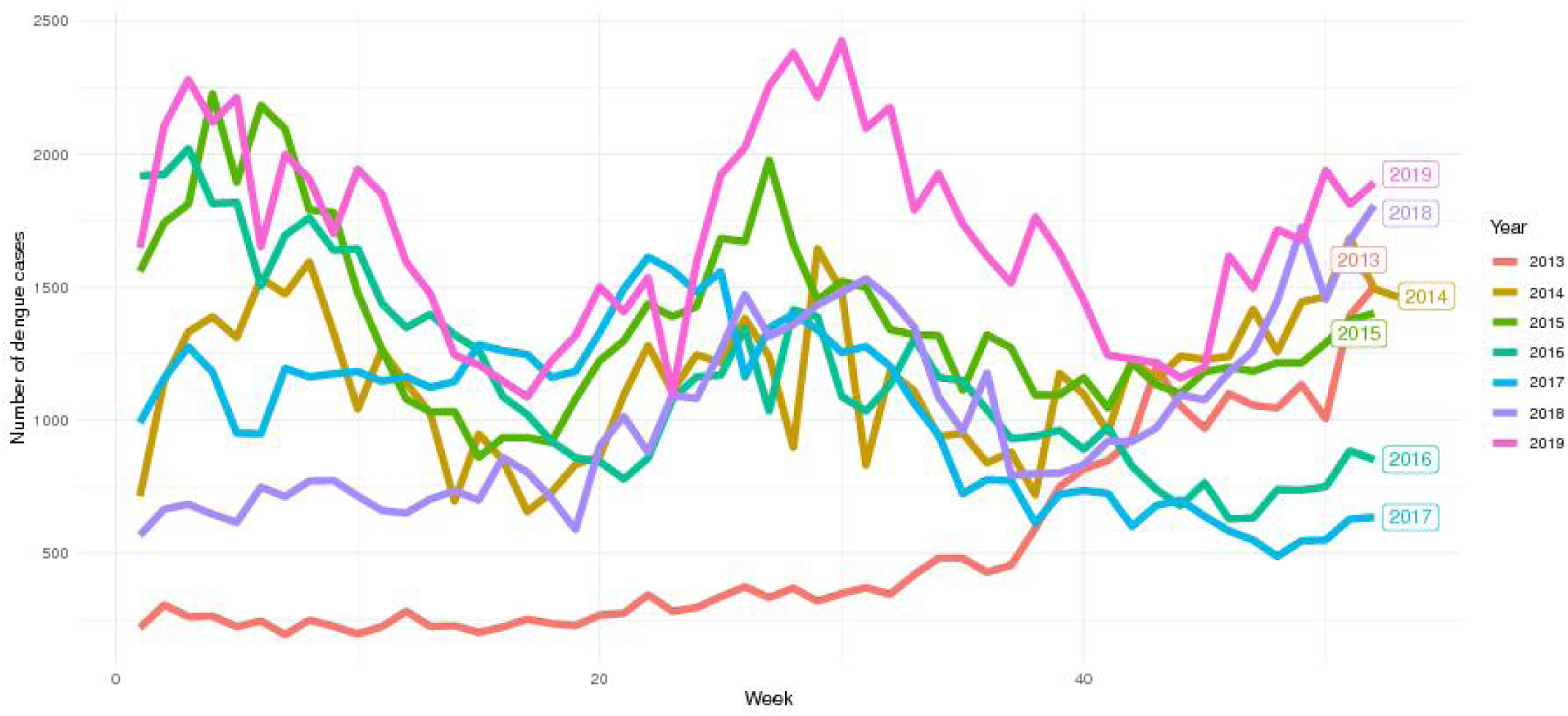
Time series plots of dengue hospitalisations by year.

#### 2.3.2 Temporal evolution and lagged effect dependency

The seasonality of dengue incidence rate (DIR) across Selangor is not as obvious as in other geographical regions in existing literature such as Thailand in Lowe et al. (2016). The weekly mean DIR peaks in the winter and finds another peak in the summer (Fig. 2).

**Figure 2:**
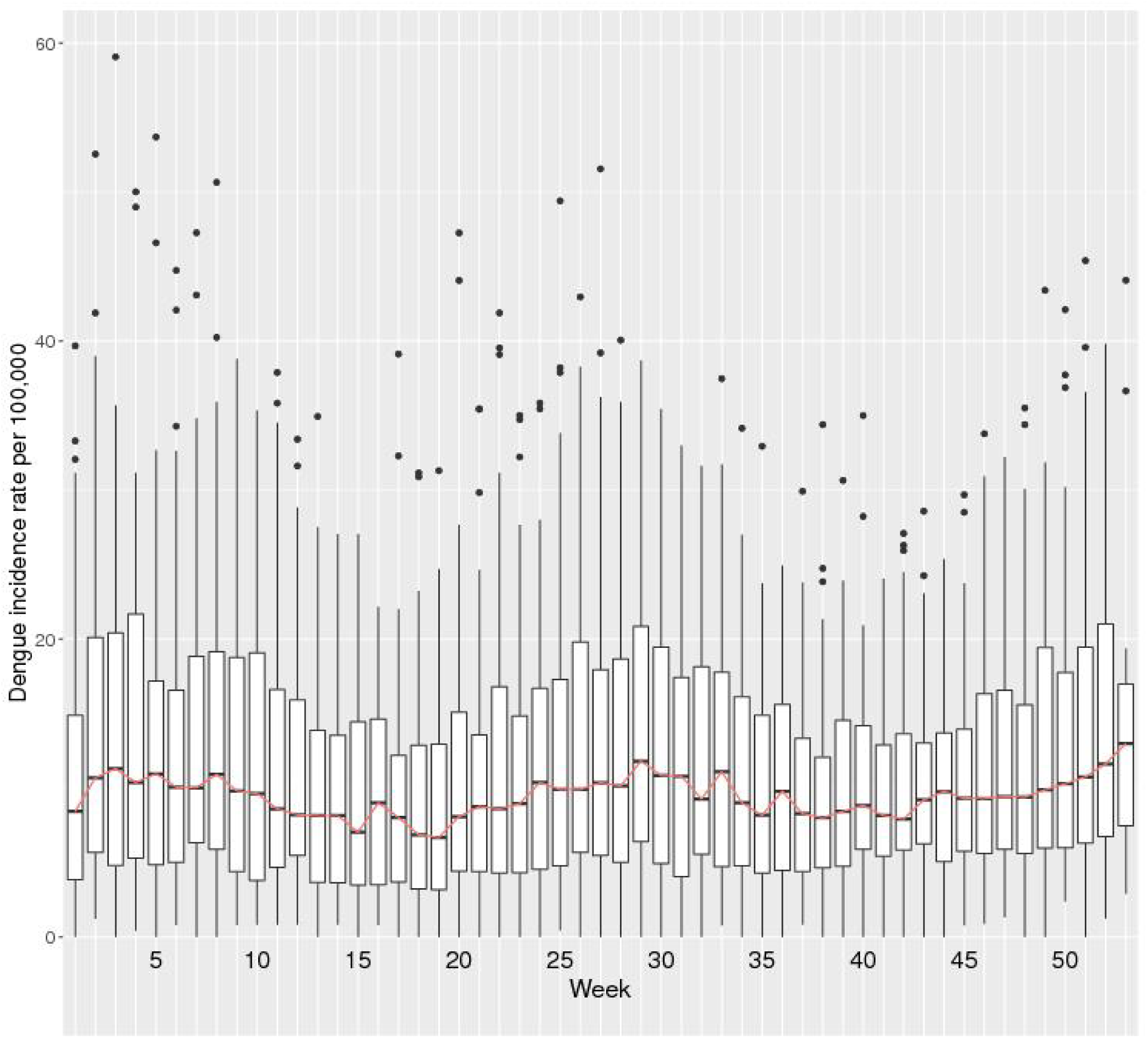
Boxplots of weekly dengue hospitalisations in the Central Region, Malaysia, 2013-2019.

Preceded by a weak El Niño events in 2014, the unusual 2015-2016 El Niño was one of the strongest El Niño in history (Lian et al., 2017). The DIR is closely related to this upward trend of both Niño1+2 and Niño4 indices during El Niño (Fig. 3). Taking out the effect of Niño4, the partial correlation between DIR and Niño1+2 index is 0.0578 only although the Niño1+2 and Niño4 are highly correlated (Fig. 4). These indices, refer to distant regions in the central/eastern equatorial Pacific, can be regarded as leading indicators of Peninsular Malaysia local climate.

**Figure 3:**
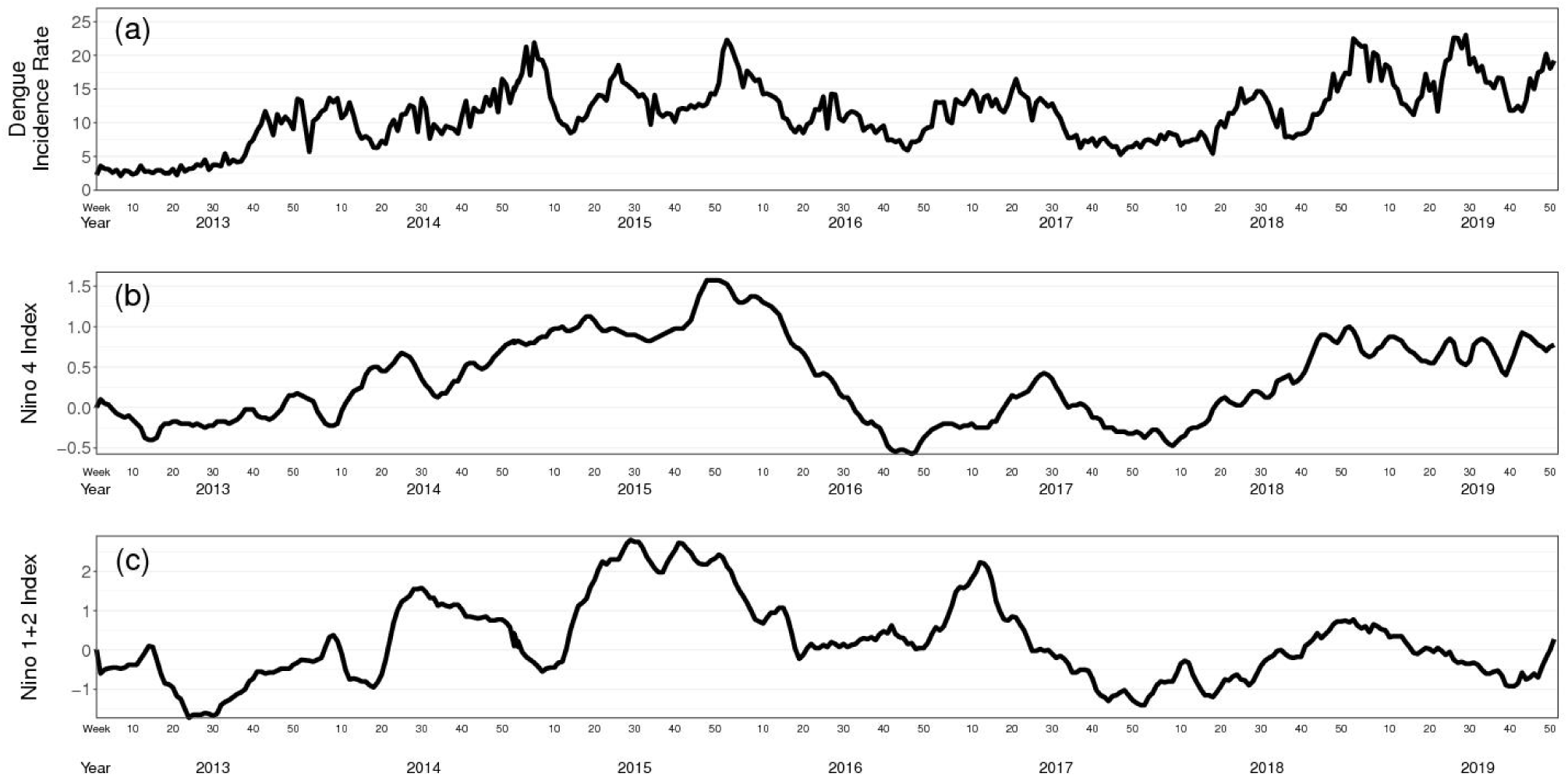
Time series of **(a)** average DIR, **(b)** lagged four-week average of Niño4 index, **(c)** lagged four-week average of Niño1+2 index in the Central Region, Malaysia for the period 2013-2019.

**Figure 4:**
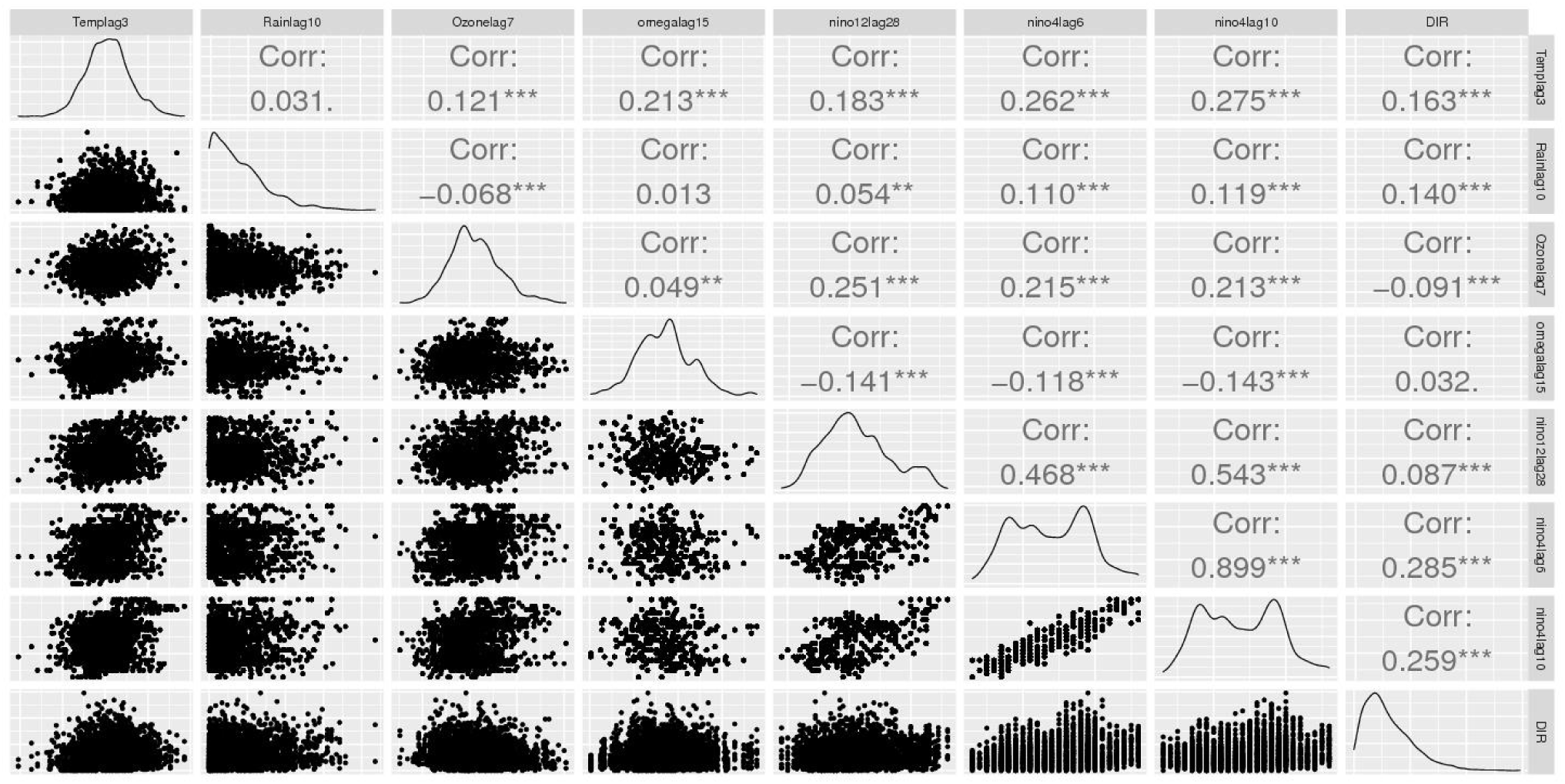
Pairwise scatter plots of the DIR along with the covariates used in the models.

Following Cheong et al. (2013), a distributed lag nonlinear model (DLNM; Gasparrini et al., 2009 and Gasparrini, 2011) is used as an exploratory tool. The dataset is aggregated into multiple single region time-series of dengue hospitalisations and environmental variables to evaluate the lag time with quartic B-splines for the predictors and lag stratifications. The relative risk (RR) at 90% quantile of temperature, ozone, rainfall, ozone, omega, Niño1+2 and Niño4 reach their maximum at a lag of 1, 10, 7, 15, 28, and 6 weeks respectively (Fig. 5). Capturing the effect of La Niña, for Niño4, the RR at 10% quantile reaches a local minimum at lag of 10 weeks. The formation of ozone is heavily influenced by sunlight and temperature (Ghazali et al., 2010). Since high temperature and presence of sunlight are the confounding factors, ozone has a strong immediate effect on dengue (Fig. 5c). The incremental cumulative RRs of rainfall has a monotonic increasing trend and have a longrange dependency throughout a long lag time. Fig. 5b shows a different pattern of short-term drought and wet scenario, with a very strong and immediate effect during drought (lag time 0-5) and a more delayed association with wet weather peak at a lag of 7 weeks. On the other hand, Fig. 5d shows a strong positive impact from the vertical velocity with a lag time of 9 weeks. It is understood that rainfall and vertical velocity are related to ground-level hydrology. A possible explanation is that drought makes people store water (Gagnon et al., 2001). Pontes et al. (2000) also suggest that household storage of water during the drought is correlated with the increase of `Aedes aegypti` vector abundance.

**Figure 5:**
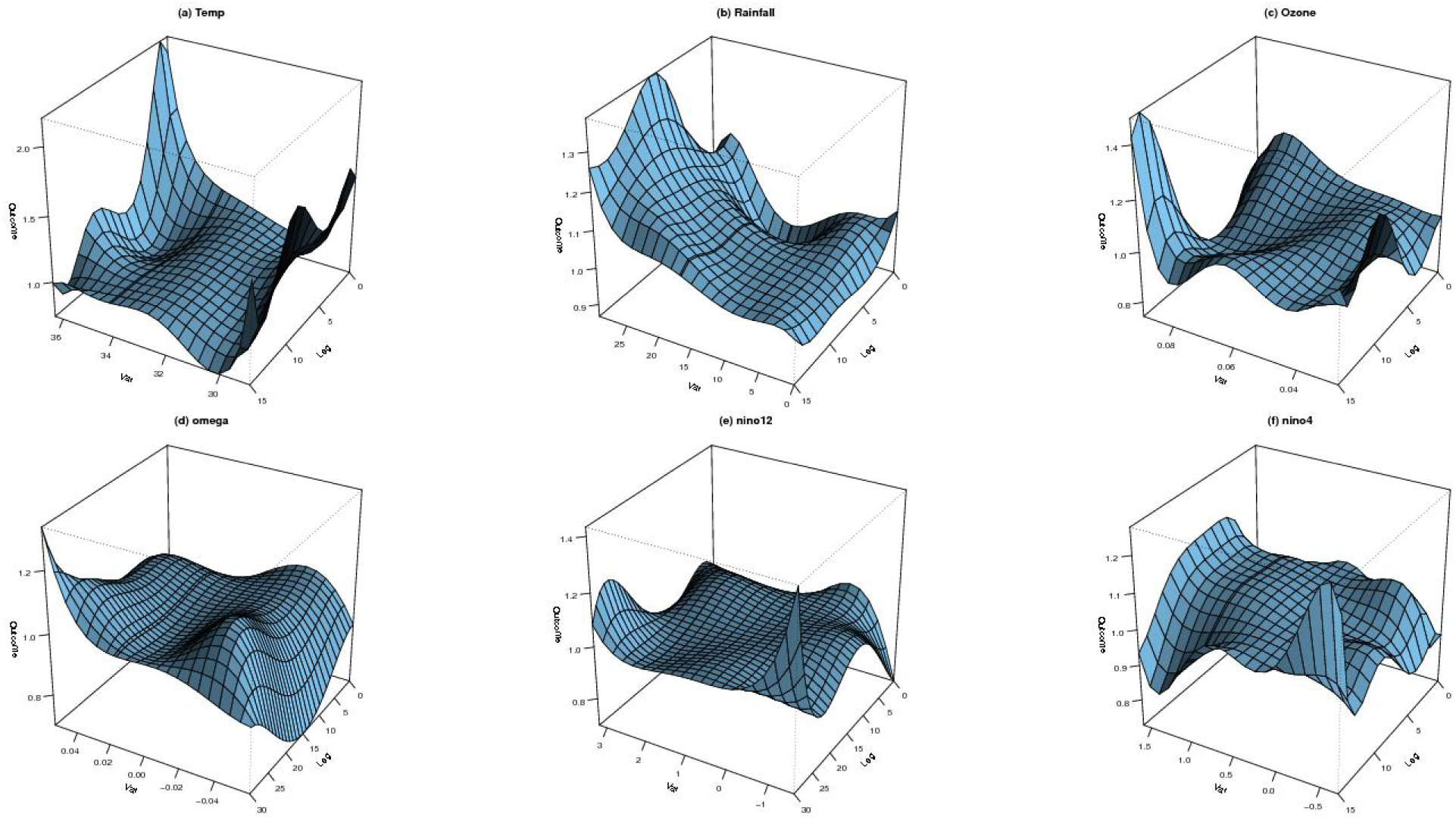
RR surface of dengue hospitalisations by six variables. The variable *Temp* is the temperature, *Rain* is total precipitation, Ozone is the ground-level ozone concentration level, omega is vertical velocity of air motion derived from omega equation, *nino12* is the Niño1+2 index, *nino4* is the Niño4 index.

#### 2.3.3 Regional variations

The Central Region area, especially districts adjacent to Kuala Lumpur, becomes hyperendemic for dengue transmission due to years of neglect (Ahmad Meer et al., 2018). Fig. 6 plots a map of DIR, temperature, rainfall level, ground-level ozone concentration level from 2013 to 2019. The districts of Gombak, Petaling, Klang and Hulu Langat generally recorded higher DIR, mean temperature and ozone concentration level compared to other districts. However, the capital city Kuala Lumpur has shown significant lower cases although among the wettest in the region. This regional variation is regarded as a function of degree of urbanisation. An explicit formulation of this type of function is generally infeasible (Chandler, 2005). An entomological explanation to this variation is related to the abundance of the breeding areas of `Aedes aegypti` and `Aedes albopictus`. In an entomological surveillance study for two villages in Selangor, Noor et al. (2018) show that two species are indoor and outdoor breeders respectively. The transmission of the dengue vector is a combined effect of two species. Hence, this socio-economic difference between districts is a source of the step change in the cases count.

**Figure 6:**
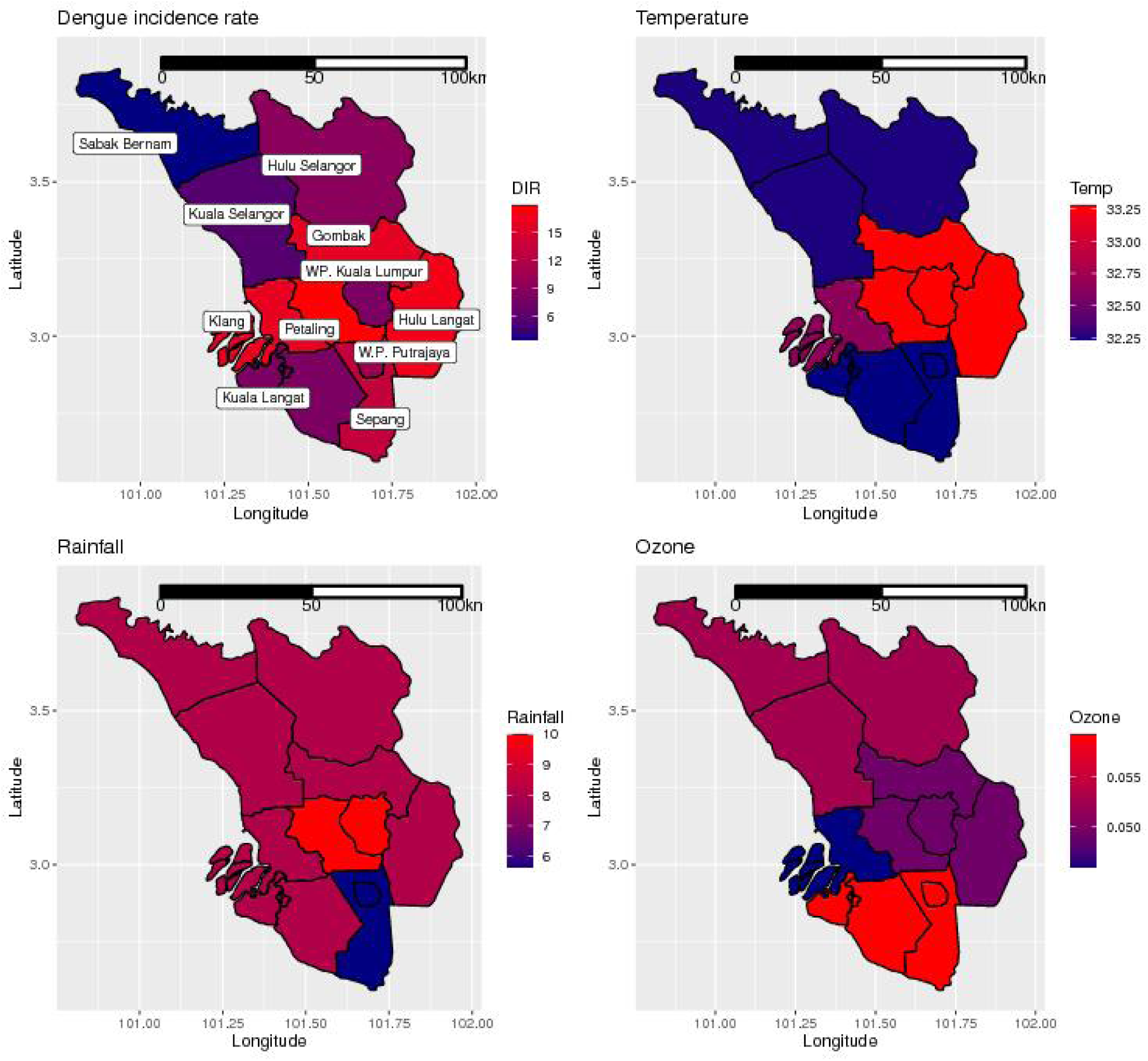
Mean weekly DIR, temperature, rainfall, ground-level ozone concentration level from 2013 to 2019 by district.

Both federal territories Kuala Lumpur and Putrajaya have lower cases than the surrounding districts. It might be attributed to the increased activity of the enforcement agencies and anti-dengue campaigns conducted in the capital city (Hassan et al., 2012).

#### 2.3.4 Spatial dependency

Without explicit spatial and temporal dependent terms, consider a Poisson generalised linear model for the disease count *Y*_*kt*_ defined in Section 2.2 of the form:

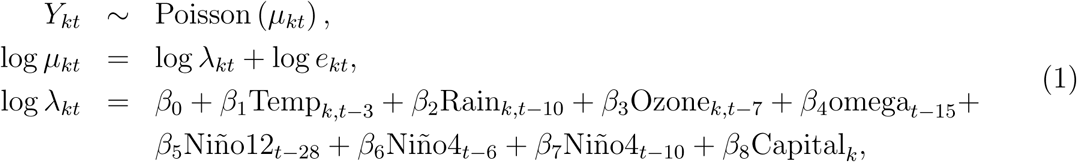

where *Y*_*kt*_ is the expected number of cases in the district *k* at time *t, e*_*kt*_ is the population size in the district *k* at time *t*, Niño12_*t*_ is the Niño1+2 index at time *t*, Niño4_*t*_ is the Niño4 index at time *t*, Ozone_*kt*_ is the ground-level ozone concentration level at time *t* in district *k*, Capital_*k*_ is a binary variable indicates whether the district is Kuala Lumpur, Temp_*kt*_ and Rain_*kt*_ is the temperature and total precipitation in the week *t*, omega_*t*_ is vertical velocity of air motion derived from weather model at time *t*. Note that here a lag of three weeks is used for temperature as one week is not a practical lag time for an EWS. We calculate the associate Moran’s I statistic (Moran, 1950) for the spatial neighbourhood matrices. The statistic is 0.7075 with a *p*-value of 0.001%. It indicates the spatial variation has not adequately been captured through the generalised linear model.

#### 2.3.5 Overdispersion

Overdispersion behaviour (Lawless, 1987) often exists in many disease count datasets. It is suggested that a negative binomial model would nicely accommodate an extra-Poisson variation in the dengue case (Lowe et al., 2011). We fit a negative binomial model using maximum likelihood estimation through a built-in R (R Core Team, 2021) function glm.nb with Equation (1) and the estimated dispersion parameter is 2.61. The amount of overdispersion is quite high. Such a statistical property can be easily described by a person is more likely to be infected by disease through close contacts. It appears that the Poisson distribution is better suited to explain the “number of infected groups” rather than the total disease count. Represented as a compound Poisson distribution with a logarithmically distributed count per group (Quenouille, 1949), the negative binomial distribution turns out to be a reasonable model.

## 3 Model developments

The Bayesian hierarchical modelling approach is a flexible framework to describe the statistical properties in the previous Section. The components of the model formulation can be, individually specified conditional to other parameters and data. In this Section, we will go through the key components of our candidate models.

### 3.1 Negative binomial regression

To overcome overdispersion, we use the negative binomial parametrisation in which introduces *r* as a universal control parameter for overdispersion (Gelman et al., 1995).

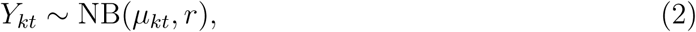

The mean and variance of the random variable are *E*[*Y*_*kt*_] = *μ*_*kt*_ and 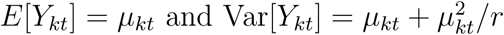 As *r* goes to infinity, the distribution of *Y*_*kt*_ converges to the Poisson distribution.

### 3.2 Besag-York-Mollié model

The Besag-York-Mollié model (BYM; Besag, York and Mollié, 1991; Besag and Kooperberg, 1995) specifies the additive relationship of the overall risk level as an intercept, the fixed effect by the covariates, the pure random effect *θ*_*kt*_ and the spatial variation component *ϕ*_*k*_:

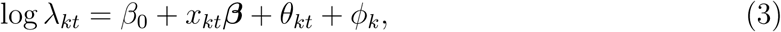

where *θ*_*kt*_ is a normally distributed unstructured error and *ϕ*_*k*_ is the structured error modelled by an intrinsic conditionally autoregressive model (ICAR). It has a conditional specification that is normally distributed with a mean equal to the average of its neighbours (*ϕ*_*k*∼*j*_) and its variance decreased as the number of neighbours *d*_*k*_ increases:

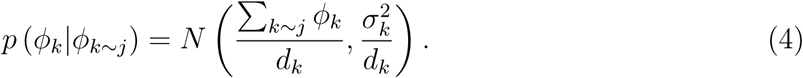

An alternative form of BYM (BYM2) model proposed by Riebler et al. (2016), Simpson et al. (2017) and Morris et al. (2019) allows a clearer dependence structure with a spatial correlation parameter ranging from a full spatial neighbourhood dependent variation and pure residual randomness in which the terms *ϕ*_*k*_ and *θ*_*kt*_ combined to one entity *ϕ*_*kt*_:

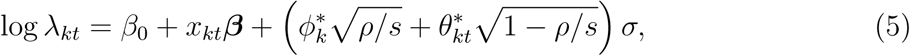

where logit (*ρ*) ∼ *N* (0, 1), 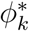 is the ICAR model, 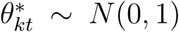, *s* is the scaling factor computed from the neighbourhood graph. Meanwhile, *σ* is the overall standard deviation of two variations.

### 3.3 Dynamic structure

Dynamic linear model (West and Harrison, 2006) enables a sequential model definition in the time series context and information propagates conditional to existing information. Taking a negative binomial model as a starting point, the Equation (2) is now a top-level observation equation, the spatio-temporal structure is defined as follows:

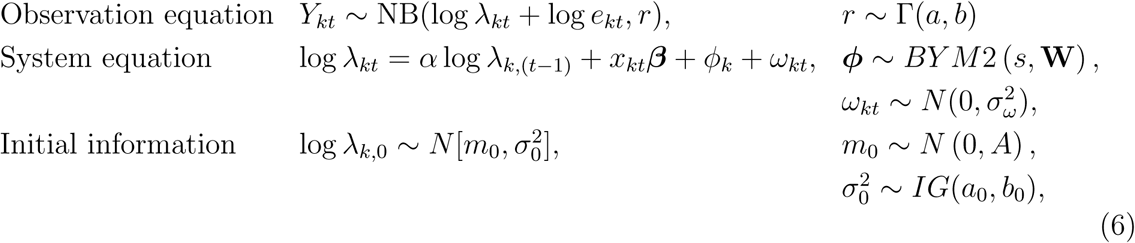

where the overdispersion parameter follows a Gamma distribution with hyperparameters *a* and *b, α* is the autoregressive (AR) parameter to control temporal dependency between adjacent time points, *ω*_*kt*_ is the Gaussian distributed evolution error. Initial information is required for this temporal structure, *s* is the scaling parameter controls the proportion of a spatial and non-spatial variation, *W* is the neighbourhood information formulated as a connected graph. The AR(1) model in the system equation could be understood as an moving average model of infinite order MA(∞) which aggregates all its lagged unexplained residuals as an additional piece of information. Sahu et al. (2009) impute the initial mean by the observed grand mean for a spatial point reference modelling problem. Alternatively, we choose to estimate the initial mean and set log *λ*_*k*0_ to follow a normal distribution with a non-informative prior for both *m*_0_ and 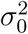.

## 4 Modelling Results

We consider five models with different levels of complexity (Table 1). The regression part of the model *x*_*kt*_***β*** is specified by the following setup:

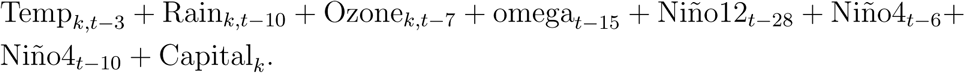

**Table 1:** Model performance by the LOO information criterion (looic), where pLOO is the estimated effective number of parameters of the model.

| Model | System equation | looic | pLOO |
| --- | --- | --- | --- |
| (A) Poisson | $\log \lambda_{kt} = \beta_0 + x_{kt}\boldsymbol{\beta}$ | $103074.7 \pm 2601.8$ | 251.3 |
| (B) NB | $\log \lambda_{kt} = \beta_0 + x_{kt}\boldsymbol{\beta}$ | $35403.1 \pm 165.4$ | 9.0 |
| (C) NB + spatial | $\log \lambda_{kt} = \beta_0 + x_{kt}\boldsymbol{\beta} + \phi_k$ | $33389.0 \pm 180.9$ | 20.6 |
| (D) NB + dynamic | $\log \lambda_{kt} = \alpha \log \lambda_{k,(t-1)} + x_{kt}\boldsymbol{\beta}$ | $29180.6 \pm 181.6$ | 1053.7 |
| (E) NB + spatial + dynamic | $\log \lambda_{kt} = \alpha \log \lambda_{k,(t-1)} + x_{kt}\boldsymbol{\beta} + \phi_k$ | $29180.9 \pm 182.5$ | 1056.6 |

No-U-turn sampler (NUTS) is used for Markov chain Monte Carlo (MCMC) sampling Hoffman et al. (2014). Nishio and Arakawa (2019) suggest that NUTS performance is better than Gibbs sampling due to the high effective sample sizes and low autocorrelations in some statistical applications.

### 4.1 Model assessment

Expected log pointwise predictive density (elpd; Vehtari et al., 2017) is used to compare model performance. The criterion is estimated by leave-one-out cross-validation to mimic out-of-sample prediction data and the looic 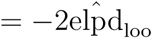 will be used for reporting to provide typical Bayesian conventional scale of deviance information criterion (DIC; Spiegelhalter et al., 2002). The overall fit of each model is summarised in Table 1. The negative binomial family of models (Model B, C, D, E) outperform the Poisson model (Model A). The negative binomial dynamic model (Model D) with the lowest looic fits the data better than the other four models. The looic of the negative binomial spatial dynamic model (Model E) and Model D differ by within one standard error.

The Model B and Model C are well-specified because the effective number of parameters (pLOO; Vehtari et al., 2017) is smaller than the actual total number of parameters in the models whilst Model A is misspecified due to failure to capture overdispersion. Bürkner et al. (2020) show that elpd/looic estimates are overly optimistic because the future observation has an influence to predictions of the past. Since the pLOO is the difference between elpd and the non-cross-validated log posterior predictive density, thus the pLOO is overestimated under any dynamic setting. The evidence is inconclusive to determine whether Model D and E are well-specified or not. A further model validation procedure is required to check their validity.

### 4.2 Environmental and regional risk factors

Although models A, B and C possess lower looic value, compared to dynamic models, they preserve a considerable explanatory power. Taking a closer look at the coefficient estimates of Model C, the coefficient estimates in the form of relative risk (RR) is shown in Table 2. The covariate lag (Niño4, 6) and lag (Niño4, 10) have a strong positive relationship with the disease, for each degree increase of the indices, the RRs increase by 46.87% and 8.44% respectively. Meanwhile, the Niño1+2 index of lag time 28 weeks decreases by 2.26% for each degree increase. The pollutant ozone has a negative effect on the disease. For every 10 parts per billion increase in concentration level, there is 3.13% decrease in dengue incidence. Kuala Lumpur has 40.40% expected cases lower than other regions. The local weather-related variables have a lesser impact on the RR with only 0.90% and −3.83% for a unit change in rainfall and temperature. The vertical velocity has a mild impact with only 3.61% RR increment for each 0.01 unit increase. The Niño4 index is the dominant factor and a negative temperature effect is seen as an adjustment to ENSO’s impact. With a positive Niño4 and a negative Niño1+2 RR, although of different lag times, this is a shred of indirect evidence that the central equatorial ENSO exerts a stronger impact on dengue disease than the convention ENSO.

**Table 2:** Parameter estimates of RR for the negative binomial spatial model (Model C)

|  | RR | Credible Interval |  |
| --- | --- | --- | --- |
|  |  | 2.5% | 97.5% |
| lag (Temp, 3) | 0.9617 | 0.9471 | 0.9761 |
| lag (Rain, 10) | 1.0090 | 1.0067 | 1.0114 |
| lag (Ozone, 7) (10ppb) | 0.9687 | 0.9561 | 0.9813 |
| lag (omega, 15) ($0.01\text{Pa s}^{-1}$) | 1.0361 | 1.0267 | 1.0456 |
| lag (Niño12, 28) | 0.9774 | 0.9602 | 0.9947 |
| lag (Niño4, 6) | 1.4687 | 1.3698 | 1.5733 |
| lag (Niño4, 10) | 1.0844 | 1.0081 | 1.1608 |
| Capital | 0.5960 | 0.1839 | 1.9244 |

### 4.3 Prediction for dengue epidemics and an early warning system

Four model validation criteria: root mean square error (RMSE), mean absolute error (MAE), continuous ranked probability score (CRPS; Hersbach, 2000), coverage at 95% nominal level (CVG; Sahu, 2021) are used for comparing out-of-sample model performance. The first two criteria evaluate the model performance in terms of mean response. The latter two are related to probabilistic forecasts. CRPS measures the discrepancy between the observations and the whole predictive distributions whilst the CVG detects underfitting and overfitting if the criterion drifts away to the nominal coverage probability of 95%. For the criteria RMSE, MAE and CRPS, better predictions correspond to their corresponding lower values. All these criteria values are calculated using the R package bmstdr developed by Sahu (2021).

Atmospheric model high resolution (HRES) provided by European Centre for Medium-Range Weather Forecasts (ECMWF) generates up to 10 days forecast (Owens and Hewson, 2018). In other words, replacing the daily mean temperature at lag time of 3 weeks by the forecasts provided by ECMWF, an EWS will have a capacity to produce outbreak detection signals at least four weeks in advance. One of the useful ways to disseminate outbreak detection is to use a visualisation called epidemic channel (Runge-Ranzinger, 2016). Once the predictive probability with a threshold level between 0.08 − 0.2 (Bowman et al., 2016) for future dengue cases exceeds a certain alarm value (e.g.: cases above than two standard deviation of the five-year average), an alarm signal forms when the weekly case numbers enter the “alarm zone”.

An out-of-sample probability forecast for the weekly reported cases in 11 districts and federal territories in the first four weeks in 2019 is generated from all model candidates. Table 3 summarises the values of the four model validation criteria from the fitted models using 2013-2018 data. Model D is the best model in terms of RMSE and MAE. Model E, although not being optimal in the first three criteria, it appears to be the most adequate model if we consider its CVG. The sensitivity and specificity (Bowman et al., 2016; Lowe et al., 2016) represent the hit rate and true negative rate of an EWS. From a disease surveillance point a view, a single miss of a disease outbreak is costly, in order to achieve the goal of identifying potential outbreaks with high sensitivity, the probability threshold level is set to a relatively small value. Setting the probability threshold level to 0.15, it means the posterior predictive distribution at 85 percentile exceeds the predefined alarm values of the reported cases greater than two standard deviations of the five-year average at each district driving an alarm signal. Using the same out-of-sample probability estimates for model validation statistics, Model E exhibits the highest sensitivity and a moderate specificity. A careful look at both Model D and E shows that the differences among the models with regard to the looic and model validation criteria are quite small. Model E appears to be a more preferable model after evaluating the overall performance measures.

**Table 3:** Model validation statistics and performance measures of early warning system derived from five models, A-E

| Model | RMSE | MAE | CRPS | CVG | Sensitivity | Specificity |
| --- | --- | --- | --- | --- | --- | --- |
| (A) Poisson | 222.63 | 203.62 | 7.97 | 2.27% | 52.94% | 44.44% |
| (B) NB | 151.86 | 91.88 | 40.62 | 81.82% | 52.94% | 51.85% |
| (C) NB + spatial | 135.81 | 84.50 | 32.69 | 72.73% | 5.88% | 88.89% |
| (D) NB + dynamic | 44.72 | 28.92 | 29.08 | 100.00% | 94.12% | 62.96% |
| (E) NB + spatial + dynamic | 51.84 | 33.44 | 38.03 | 95.45% | 94.42% | 70.37% |

We found that the most important RR comes from Niño4. It makes a longer-term prediction of dengue outbreaks feasible. Meng et al. (2020) shows that a complexity-based approach allows us to forecast the magnitude of an ENSO event one year in advance. Ham et al. (2019) utilise a convolution neural network (CNN) to predict zonal SST (in their example, Niño3.4 region) by learning from historical simulations of a multi-model ensemble (Bellenger et al., 2014). Ballpark figures generated from a simpler EWS with ENSO information can be then assessed by the government agency. A longer-term climate uncertainty analysis (Yip et al., 2011; Northrop and Chandler, 2014) can be easily plugged into a disease mapping setting (e.g.: Baker et al., 2021).

## 5 Discussion

This paper presents a Bayesian spatio-temporal modelling framework leading to a full implementation of an EWS for dengue outbreaks from upstream data source to production. We propose to utilise some global and regional climate observation and variables derived from reanalysis data for a more accurate forecast. Exploratory data analysis methods show a long range of lag times is required for some synoptic scale meteorological variables namely Niño12 and Niño4 indices. Our proposed holistic assessment goes beyond a single cross-validation metric. The whole assessment consists of calculation of out-of-sample predictive accuracy in multiple ways and alert signal evaluation. The methodology developed in this study can potentially be used to build a similar EWS in other countries or regions in Malaysia.

Dissimilar to horizontal wind, the vertical motion is often neglected due to its unobserved nature. In contrast to the environmental variables considered in previous studies, this study also considers a vertical velocity of air motion derived from reanalysis data and reveals to have a mild effect to the epidemics. Similar to many other studies, temperature and rainfall are used in the regression formula. Contrary to the findings of other studies (e.g. Lowe et al., 2016), the estimated coefficient of lagged temperature is negative. This appears to be a case of a local adjustment to a larger scale regional effect dominated by ENSO. The estimate of lagged ozone ties well with the biological argument based on Aedes’s gonotrophic cycle in Wong et al. (2011).

The RR estimates from the Model C exhibit that strong lagged anomalous warming in the Niño4 region has a strong positive effect on dengue hospitalisations. Consistent with our present findings, Gagnon et al. (2001) also report a significant positive correlation between El Niño and dengue epidemics in multiple countries. With a less-than-one RR for the lagged Niño1+2 index, cooling in the eastern tropical Pacific contributes to the increased dengue. This distinct relationship suggests that both El Niño and El Niño Modoki play a role in the epidemics. A previous study by Petrova et al. (2019) mentions that dengue epidemics can be associated with different teleconnections for different time lags. Dengue transmission is sensitive to the variability of rainfall due to its cumulative nature. A recent finding shows that a strong positive IOD which leads to drought (Amirudin et al., 2020) can be linked with the pre-existing El Niño Modoki with lead time up to one year (Doi et al., 2020). Our results align to these claims.

Due to data limitations, the impact from spatio-temporal variations of virus serotype are missing from the study. An anomalous upsurge happens twice in our study period, the first one occurred in the 2013 summer is verified by microbiology evidence (Ng et al., 2015). The second one observed in early 2019 is thought to be due to another serotype shift. A self-service EWS received a user feedback that change of predominant serotype alone attains a 50% of sensitivity of outbreak detection (Hussain-Alkhateeb et al., 2018).

Although the transmission dynamics is proven to be temperature-dependent (e.g.: Chen et al., 2012), the relationship between entomological parameters and the environment variables has not yet been clearly studied. A recent article by Sun et al. (2021) study a residential-block-level dengue vector population in Singapore. It is shown that the `Aedes` abundance is heavily associated with the building age and managed vegetation cover. With modern geographic information systems (GIS) technology, these information can be incorporated in the future work.

Thanks to the flexibility of the modelling framework in this research article, joint modelling on multiple diseases is a possible methodological extension. Caminade et al. (2017) show the mosquito-borne transmission of Zika in South America is fueled by the El Niño climate phenomenon. Funk et al. (2016) suggest, with their extensive sensitivity analysis, models for dengue transmission can be useful for handling the dynamics of Zika transmission. Held et al. (2005) demonstrate that the joint modelling approach on multiple diseases achieves a gain in precision of the RR estimates. Niriella et al. (2021) spot a sharp decrease in dengue cases for the second quarter of 2020 compared with pre-COVID-19 peaks in Sri Lanka. The drastic measures imposed by the Sri Lanka government regarding COVID-19 outbreaks help the reduction of hospitalisations. An identical pattern is also found during the first five phrases COVID-19 lockdown in Malaysia (Ong et al., 2021). A vector autoregression component (VAR; Spencer, 1993) can be added to our current setup to incorporate lagged effect dynamically from other variables. Implemented in Stan language (Carpenter et al., 2017), conditional dependence such as spatial heterogeneity, temporal dynamics and covariate structure can be simply introduced and modified under the hierarchical Bayesian modelling paradigm, allowing for greater modelling flexibility.

## 6 Conclusion

The SST anomalies with a lag time of six weeks in the central equatorial Pacific region is the most crucial driver to the Central Region of Malaysia dengue hospitalisations. The EWS built on a Bayesian spatio-temporal hierarchical model yields reliable forecasts to help out dengue disease outbreak surveillance for at least four weeks in advance.

## Data Availability

The data that support the findings of this study are available upon request from the authors.

## Acknowledgments

The authors thank Dr Edith Cheng for helpful comments. We gratefully acknowledge the Vector Borne Disease Sector, Disease Control Division, Ministry of Health of Malaysia, Department of Statistics of Malaysia, Department of Environment and Malaysian Meteoro-logical Department for providing the information and dengue data used in this study.

